# Genome-wide analyses of 200,453 individuals yields new insights into the causes and consequences of clonal hematopoiesis

**DOI:** 10.1101/2022.01.06.22268846

**Authors:** Siddhartha P. Kar, Pedro M. Quiros, Muxin Gu, Tao Jiang, Ryan Langdon, Vivek Iyer, Clea Barcena, M.S. Vijayabaskar, Margarete A. Fabre, Paul Carter, Stephen Burgess, George S. Vassiliou

## Abstract

Clonal hematopoiesis (CH) is one of the most extensively studied somatic mutational phenomena, yet its causes and consequences remain poorly understood. We identify 10,924 individuals with CH amongst 200,453 whole-exome sequenced UK Biobank participants and use their linked genome-wide DNA genotypes to map the landscape of inherited predisposition to CH. We increase the number of European-ancestry genome-wide significant (*P*<5×10^−8^) germline associations with CH from four to 14 and identify one new transcriptome-wide significant (*P*<3.2×10^−6^) association. Genes at new loci implicate DNA damage repair (*PARP1, ATM*, and *CHEK2*), hematopoietic stem cell migration/homing (*CD164*), and myeloid oncogenesis (*SETBP1*) in CH development. Several associations were CH-subtype specific and, strikingly, variants at *TCL1A* and *CD164* had opposite associations with *DNMT3A*-versus *TET2*-mutant CH, mirroring recently reported differences in lifelong behavior of these two most common CH subtypes and proposing important roles for these loci in CH pathogenesis. Using Mendelian randomization, we show, amongst other findings, that smoking and longer leukocyte telomere length are causal risk factors for CH and demonstrate that genetic predisposition to CH increases risks of myeloproliferative neoplasia, several non-hematological malignancies, atrial fibrillation, and blood epigenetic age acceleration.

## Introduction

The pervasive effects of ageing and somatic mutation shape the landscape of human disease in later life^1^. A ubiquitous feature of ageing is the development of somatic mutation-driven clonal expansions in aged tissues^2,3^. In blood, somatic mutations that enhance cellular fitness of individual hematopoietic stem cells (HSCs) and their progeny, give rise to the common age-related phenomenon of clonal hematopoiesis (CH)^4–7^. CH becomes increasingly prevalent with advancing age^4–6^ and is associated with an increased risk of hematological cancers^4,5,8,9^ and of some non-hematological conditions^5,10,11^. However, our understanding of the biological basis for these associations remains limited, as does our ability to explain how CH driver mutations promote clonal expansion of mutant HSCs^12^. In fact, whilst CH is defined by its association with somatic mutations, its development is influenced by non-mutation factors^13–16^ and by the heritable genome^17,18^, in ways that remain poorly understood.

Insights into the causes and consequences of CH are confounded by its intimate relationship with ageing. Moreover, even when robust associations are identified, their causality can be difficult to establish. Here, we perform a comprehensive investigation of the genetic and phenotypic associations of CH in 200,453 United Kingdom Biobank (UKB) participants, yielding a step change in our understanding of CH pathogenesis. Our study reveals multiple new germline loci associated with CH, including several that interact with specific CH subtypes, uncovers causal links between CH and diverse pathological states across organ systems, and provides evidence for causal associations between smoking and telomere length and CH risk, amongst a series of novel insights.

## Results

### Prevalence of CH and its distribution by age and sex in the UKB

To identify individuals with CH, we analyzed blood whole exome sequencing (WES) data from 200,453 UKB participants^19^ aged 38-72 years (Extended Data Fig. 1a-c). We called somatic mutations in 43 CH genes (Supplementary Table 1) and filtered these against a predefined list of CH driver variants (Supplementary Tables 2 and 3). This identified 11,697 mutations (Supplementary Table 4) in 10,924 individuals (UKB prevalence: 5.45%). *DNMT3A, TET2*, and *ASXL1* were most commonly involved (79% of all mutations), followed by mutations in DNA damage response genes *PPM1D, TP53*, and *ATM*; splicing factor genes *SRSF2* and *SF3B1*; *JAK2* and *GNB1* (Fig. 1a), in line with previous reports^4,5,17^. Most CH carriers (n=10,228) harbored one and some (n=696) 2-4 mutations; most of which were missense variants dominated by cytosine-to-thymine (C>T) transitions (Extended Data Fig. 1d-f). The mean variant allele fraction (VAF) was 0.12 and VAF distribution did not differ between mutation types (Extended Data Fig. 1g and h). VAF distribution did differ between individual genes (Fig. 1a), although some of this variation was probably influenced by variation in sequencing depth (Supplementary Table 1).

**Fig. 1:**
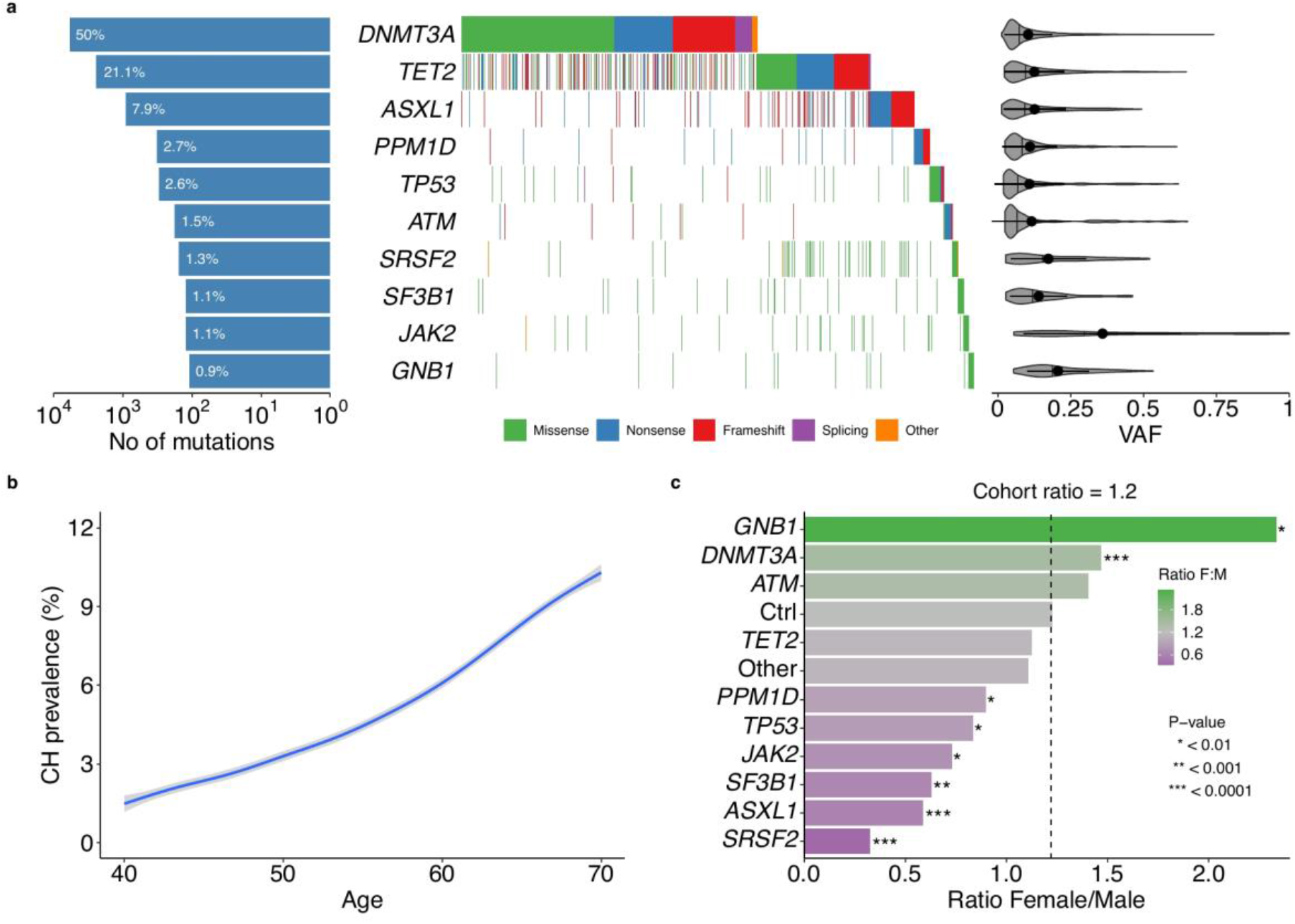
Characterization of CH in the UK Biobank. **a**, Composite plot summarizing mutations in the ten most common driver genes in 10,924 individuals with CH. Each column in the waterfall plot represents a single individual, with mutation types color-coded. Bars on the left quantify mutations per gene as a percentage of all CH mutations identified. Violin plots on the right show the distribution of variant allele fractions (VAFs), with vertical lines represent the median and dots with horizontal lines the mean ± standard deviation. **b**, Prevalence of CH in the cohort with advancing age. The blue line represents the smoothed model fitted to a generalized additive model with 95% confidence interval (CI; grey shadow**). c**, Bar plot showing the female to males (F:M) ratio of CH carriers with mutations in the ten most common driver genes. “Other” represents the remaining driver genes grouped together and “Ctrl” the ratio for individuals without CH. Dotted vertical line shows the F:M ratio observed in the full cohort (F:M=1.2). *P*-values are from a Chi-square test comparing the distribution for each gene to “Ctrl”.

CH prevalence rose progressively with age (*P*<10^−300^; Fig. 1b), as did clone size measured by VAF (*P*=8.2×10^−37^; Extended Data Fig. 2a). Females and males were similarly affected with similar median ages (Extended Data Fig. 2b). The age-related rise in prevalence differed between drivers: compared to *DNMT3A*, mutations in *ATM* were observed earlier and those in *ASXL1, PPM1D, SF3B1*, and *SRSF2* were observed later (Extended Data Fig. 2c). Furthermore, we noted significant differences in the prevalence of different CH gene mutations between sexes, with *GNB1* and *DNMT3A* mutations more frequent in females and *PPM1D, TP53, JAK2, SF3B1, ASXL1*, and *SRSF2* mutations more frequent in males (Fig. 1c), reflecting their relative prevalence in myeloid malignancies^20^.

### Associations between CH and traits/diseases prevalent at the time of blood sampling

To identify associations between CH and traits or diseases prevalent at the time of enrolment to the UKB, we performed regression analyses adjusting for age, sex, smoking status, WES batch and the first ten genetic ancestry principal components. Individuals with CH showed higher average platelet, leukocyte, reticulocyte, and neutrophil counts and red blood cell distribution width (RDW), but lower eosinophil counts (Fig. 2a). These associations were more pronounced in individuals with large CH clones (VAF≥0.1; Fig. 2a; Supplementary Table 5). *JAK2*-driven CH was associated with markedly higher platelet counts, RDW and hemoglobin/hematocrit (HGB/HT) levels. In contrast, splicing factor-mutant CH was associated with lower HGB/HT and higher mean red cell volume (MCV; Fig. 2a; Supplementary Table 5). We also found that CH status was associated with lower levels of total and low-density lipoprotein cholesterol (Fig. 2b; Supplementary Table 6). Advancing age increased the risk of CH by 6.7% per year (OR=1.07, 95%CI: 1.06-1.07, *P*<10^−300^; Fig. 2c); and CH status was associated with increased prevalence of hypertension, but not obesity or type 2 diabetes (T2D; Fig. 2c; Supplementary Table 7). Also, individuals with CH were more likely to be current, past, or “ever” smokers, an association that held true for different forms of CH and was strongest for *ASXL1-*mutant CH (Fig 2c and 2d; Supplementary Table 7).

**Fig. 2:**
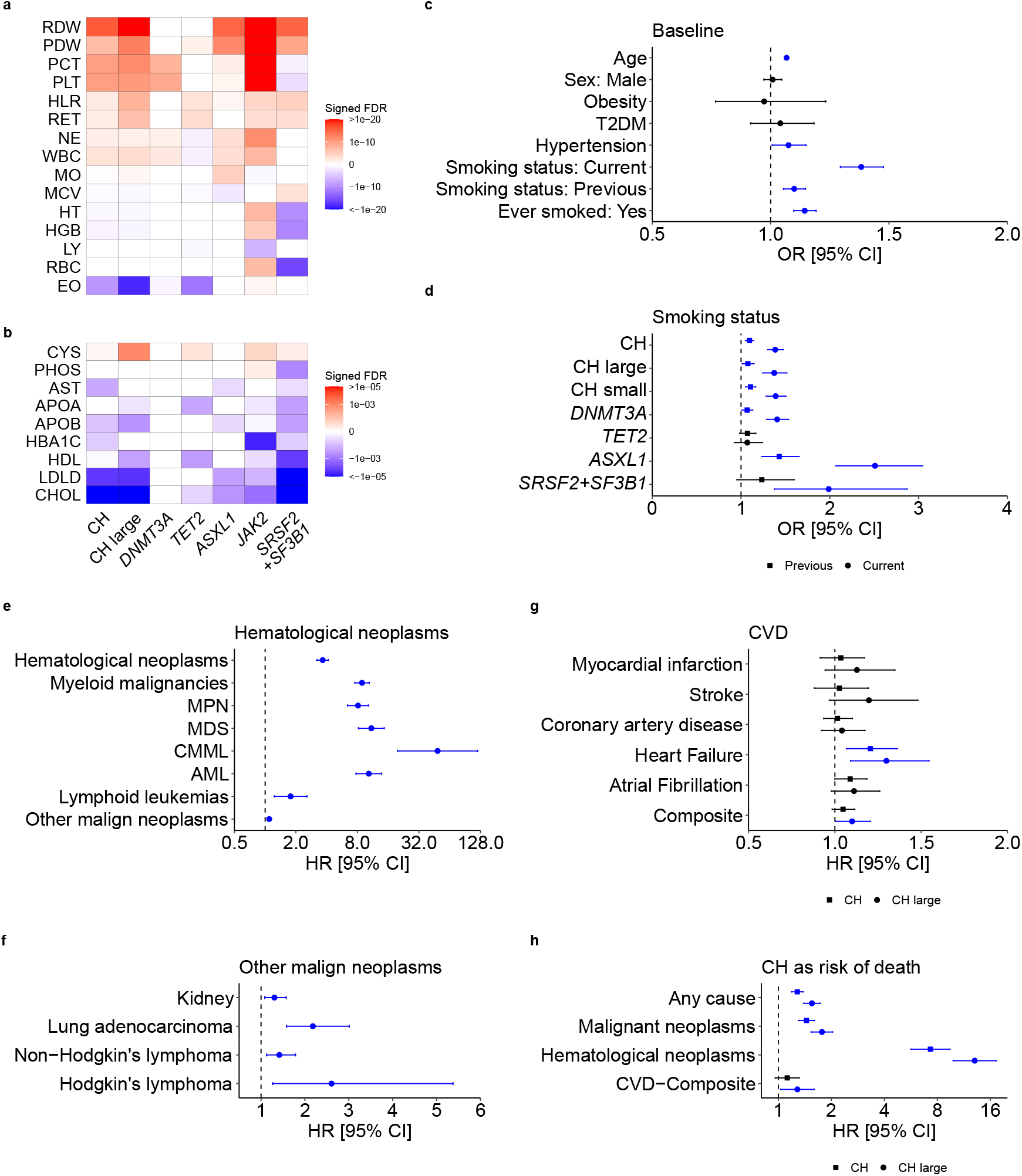
Associations between CH and diverse traits/diseases. **a-b**, Heatmaps showing associations between overall CH, CH with large clones, and CH driven by *DNMT3A, TET2, ASXL1, JAK2*, and *SRSF2*+*SF3B1* mutations and: a, blood cell counts/indices or b, biochemical analytes. Colors depict statistical significance of differences compared to individuals without CH, as signed false discovery rate (FDR) values. **c**, Forest plot showing the odds ratios (ORs) for associations between CH and selected traits/diseases prevalent in UKB participants at baseline. **d**, Forest plot showing the ORs for associations between CH subtypes and smoking status, for previous and current smokers. **e-h**, Forest plots showing the hazard ratios (HRs) for associations between CH at baseline and subsequent: **e**, hematological neoplasms, **f**, other malignant neoplasms, **g**, cardiovascular diseases, and h, selected causes of death. For g and **h**, both overall CH and CH characterized by large clones (“CH large”) are shown. ORs/HR markers with a *P*-value<0.05 are depicted in blue. Error bars represent 95% confidence intervals (CIs). Numerical values for ORs/HRs, 95% CIs, and *P*-values are reported in Supplementary Tables 5—13. Abbreviations: RDW, red blood cell (erythrocyte) distribution width; PDW, platelet distribution width; PCT, plateletcrit; PLT, platelet count: WBC, white blood cell (leukocyte) count; NE, neutrophil count; HLR, high light scatter reticulocyte count; RET, reticulocyte count; MO, monocyte count; MCV, mean corpuscular volume; HT, hematocrit percentage; HGB, hemoglobin concentration; LY, lymphocyte count; RBC, red blood cell (erythrocyte) count; EO, eosinophil count; CYS, cystatin C; PHOS, phosphate; AST, aspartate aminotransferase; HBA1C, glycosylated hemoglobin; APOA, apolipoprotein A; APOB, apolipoprotein B; HDL, HDL cholesterol; LDLD, LDL direct cholesterol; CHOL, total cholesterol; T2DM, type 2 diabetes mellitus; MPN, myeloproliferative neoplasms; MDS, myelodysplastic syndromes; AML, acute myeloid leukemia; CMML, chronic myelomonocytic leukemia.

### Associations between CH and incident disease

We next investigated relationships between CH at baseline and traits/diseases that developed subsequently (Supplementary Table 8) and identified strong associations with incident myeloid malignancies (MM) and associated sequelae (Extended Data Fig. 3a and Supplementary Table 9). The association was strong for all MM subtypes and highest for chronic myelomonocytic leukemia (CMML; Fig. 2e), whilst large clone CH increased the risk of MMs three-to-five-fold compared to small clone CH (VAF<0.1; Extended Data Fig. 3b; Supplementary Table 10). *SF3B1* and *SRSF2* mutations conferred very high risks of CMML and myelodysplastic syndromes (Extended Data Fig. 3c; Supplementary Table 10). CH was also associated with increased risks of Hodgkin’s and non-Hodgkin’s lymphomas and non-hematological neoplasia, including lung, head and neck, kidney, bladder, colorectal, and stomach cancers (Fig. 2f; Supplementary Table 11). The association of CH with lung adenocarcinoma was consistently observed across large and small clones, and with *DNMT3A* and *ASXL1* mutations, whilst the association with overall CH persisted in self-reported never-smokers (Extended Data Fig. 3d).

As CH was previously identified as a risk factor for ischemic cardiovascular disease (CVD)^5,10,21^, we examined the association in this much larger cohort (Fig. 2g; Supplementary Table 12). Using multivariable models, we did not find a significant association between CH and ischemic CVD, including coronary artery disease (CAD) and stroke; however, we did find significantly increased risks of heart failure and a composite of all CVD conditions (Fig. 2g; Supplementary Table 12). Using a bivariable model including age as the only other covariate, we also found a significant association with atrial fibrillation, with an effect size estimate consistent with that in multivariable analysis (Extended Data Fig. 3e; Supplementary Table 12). Our multivariable analyses also found significant associations between CH and increased risk of death from any cause, malignant neoplasm, and hematological neoplasm, whilst large clone CH was also associated with an increased risk of death due to the composite of CVDs (Fig. 2h; Supplementary Table 13).

### Heritability and cell type-specific enrichment of polygenic susceptibility to CH

To identify heritable determinants of CH risk, we performed a genome-wide association study (GWAS) on the 184,121 individuals with genetically inferred European ancestry to identify common (minor allele frequency (MAF)>1%) germline genetic variants predisposing to CH. In the GWAS, we compared 10,203 individuals with CH with 173,918 individuals without CH, after quality control of the germline genotype data. Linkage disequilibrium score regression (LDSC)^22^ showed little evidence of inflation in test statistics due to population structure, with an intercept of 1.009 and lambda genomic control factor of 0.999. The narrow-sense (additive) heritability of CH was estimated at 3.57% (s.e.=0.85%). We partitioned the heritability of CH across four major histone marks observed in 10 cell type groups aggregated from 220 cell-type specific annotations^23^ and identified strong enrichment of the polygenic CH association signal in histone marks enriched in hematopoietic cells (*P*=5.9×10^−5^; Fig. 3a; Supplementary Table 14). Next, we partitioned the heritability of CH across open chromatin state regions in various hematopoietic progenitor cells and lineages^23,24^. We found evidence of CH heritability enrichment in accessible chromatin regions in HSCs, common lymphoid and myeloid progenitors, multipotent and erythroid progenitors, and B cells (Fig. 3b; Supplementary Table 15). Overall, these findings are in keeping with the intuitive assumption that the CH GWAS exerts its greatest biological effect on HSC/progenitor populations.

**Fig. 3:**
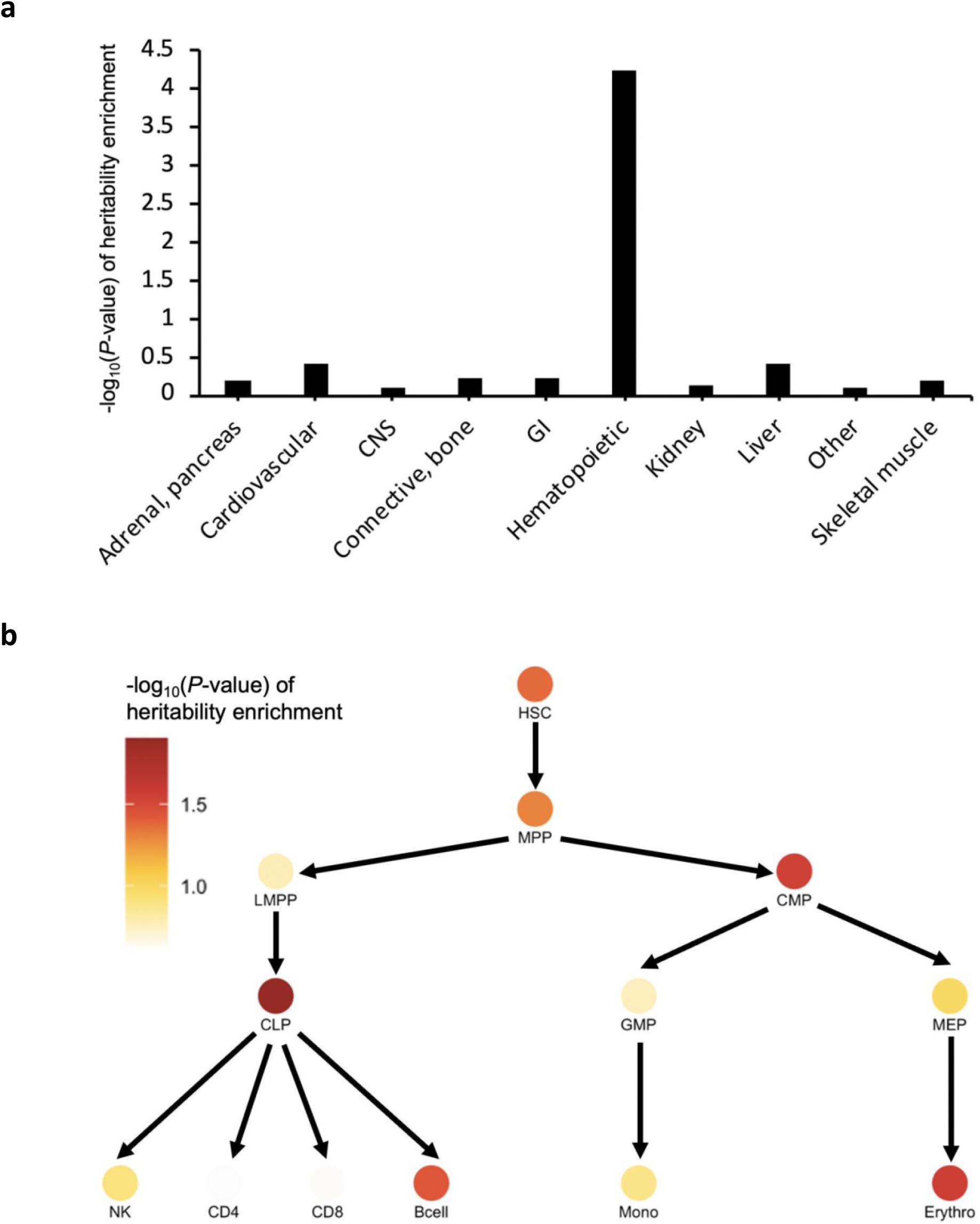
Cell type-specific enrichment of the CH polygenic signal. **a**, Heritability enrichment of CH across histone marks profiled in 10 cell type groups. **b**, Heritability enrichment of CH across open chromatin regions identified by ATAC-seq in hematopoietic progenitor cells/lineages at different stages of differentiation. Partitioned heritability cell-type group analysis in the LDSC software was used to compute these enrichments and corresponding *P*-values. The data underlying the figures is available in Supplementary Tables 14 and 15. Abbreviations: CNS, central nervous system; GI, gastrointestinal; CLP, common lymphoid progenitor; CMP, common myeloid progenitor; MPP, multipotent progenitor; HSC, hematopoietic stem cell; GMP, granulocyte/macrophage progenitor; LMPP, lymphoid-primed multipotent progenitor; NK, natural killer cell; Mono, monocyte; Erythro, erythroid progenitor; LDSC, linkage disequilibrium score regression; ATAC-seq, (Assay for Transposase-Accessible Chromatin using sequencing).

### Germline genetic loci associated with overall CH susceptibility

Linkage disequilibrium (LD)-based clumping of 9,715,652 common variants identified seven independent (*r*^2^<0.05) genome-wide significant loci (lead variant *P*<5×10^−8^) associated with risk of developing CH, including three previously reported^17^ European-ancestry CH loci: two at 5p15.33-*TERT* and one at 3q25.33-*SMC4* (Fig. 4a; Supplementary Table 16). We identified a new top variant in the 5p15.33 region, rs2853677 (*P*=2.4×10^−50^), which was weakly correlated (*r*^2^=0.19) with the previously reported^17^ top variant, rs7705526 (*P*=3.4×10^−44^ in our analysis). Overall, there was evidence for three independent (*r*^2^<0.05) signals at 5p15.33 marked by lead variants rs2853677, rs13156167, and rs2086132, the latter representing a new signal independent of the two previously published^17^ signals rs7705526 and rs13167280. After approximate conditional analysis^25^ (Supplementary Table 17) conditioning on the three lead variants in the *TERT* region, the previously published top variant, rs7705526, continued to remain genome-wide significant suggesting that it represented a fourth signal in this region. Conditional analysis also highlighted the existence of a fifth independent association at 5p15.33 marked by rs13356700 ∼776 kb from *TERT* and ∼34 kb from *EXOC3* (Supplementary Table 17) that encodes an exocyst complex component implicated in arterial thrombosis^26^. The variant rs13356700 was in strong LD (*r*^2^=0.84) with rs10072668 that is associated with HGB/HT^27^. At 3q25.33-*SMC4*, the previously reported^17^ top variant, rs1210060191, was not captured in the UKB and our top association was rs12632224 (*P*=2.3×10^−9^). We also identified three other novel genome-wide significant loci associated with overall CH susceptibility (Fig. 4a; Supplementary Table 16): 4q35.1-*ENPP6* (rs13130545), 6q21-*CD164* (rs35452836), and 11q22.3-*ATM* (rs11212666).

**Fig. 4:**
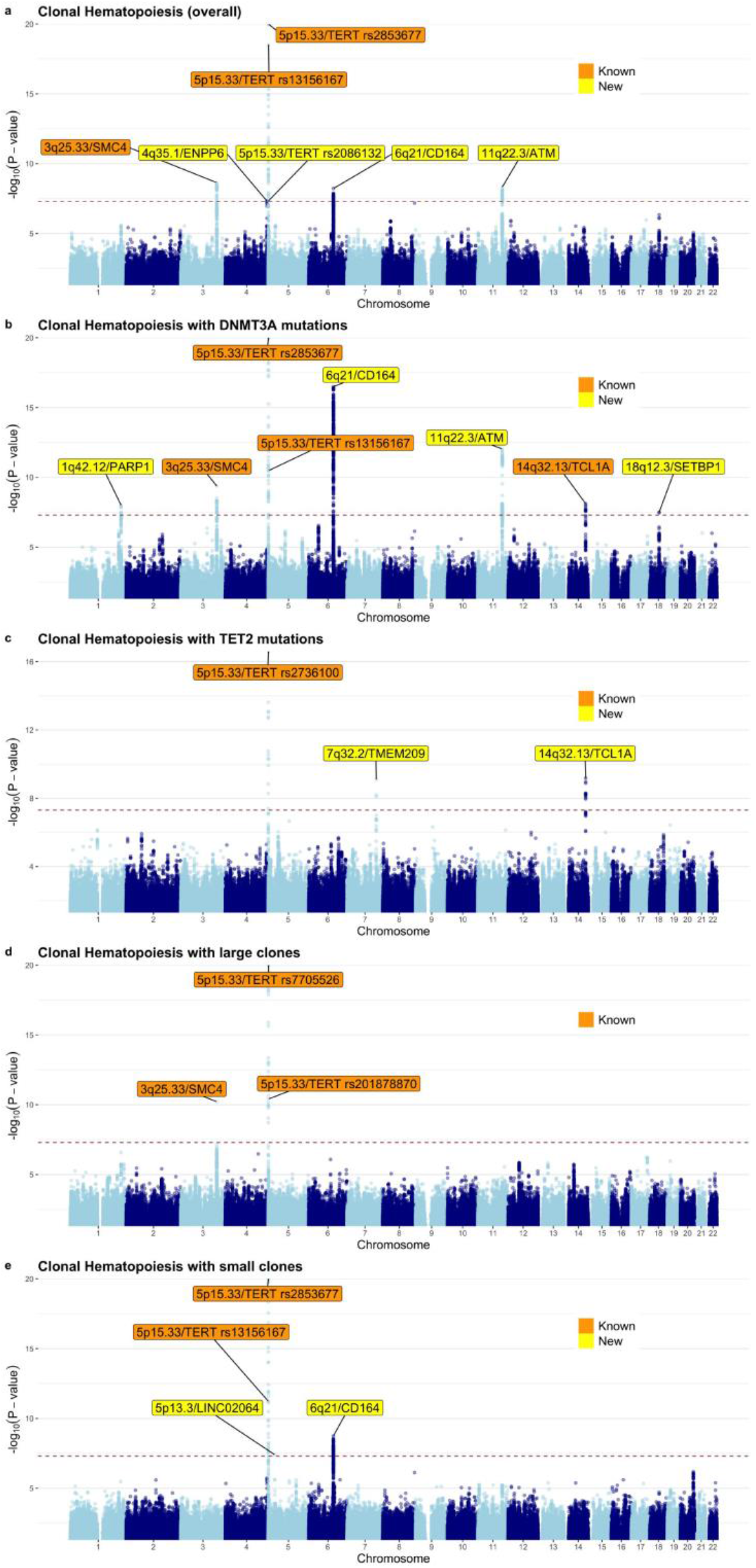
Manhattan plots displaying genome-wide associations between common germline genetic variants and each of five CH traits. The Y-axes depict *P*-values (-log_10_) for associations derived from the non-infinitesimal mixed model association test implemented in BOLT-LMM. The X-axes depict chromosomal position on build 37 of the human genome (GRCh37). The dotted lines indicate the genome-wide significance threshold of *P*=5×10^−8^. Known (previously published) and new loci are indicated by cytoband and target gene (based on the prioritization exercise described in the text). Since there were multiple independent loci at 5p15.33 (LD *r*^2^<0.05), we also label the 5p15.33 signals using the lead variant rs number for each signal. Our prioritization exercise was focused on protein coding genes near each lead variant and since there were no protein coding genes within 1 Mb of the lead variant at 5p13.3, we labeled this association using the nearest non-coding RNA. The CH traits corresponding to each Manhattan plot are: **a**, overall CH. **b**, CH with mutant *DNTM3A*. **c**, CH with mutant *TET2*. **d**, CH with large clones. **e**, CH with small clones.

### CH GWAS stratified by gene and clone size, association heterogeneity, and rare variant associations

Next, we investigated whether the development of certain CH subtypes may be affected by germline variants. Thus, we performed GWAS for four additional CH traits – stratifying by the two main mutated genes in CH, *DNMT3A* and *TET2*, and by clonal size, differentiating large and small clones. Focusing on 5,185 individuals with *DNMT3A* and 2,041 with *TET2* mutations and using the 173,918 individuals of European ancestry without detectable CH as controls, we identified eight and three genome-wide significant loci associated with *DNMT3A*- and *TET2*-mutant CH, respectively (Figs. 4b and 4c; Supplementary Tables 18 and 19). We replicated the only previously published European-ancestry CH risk locus associated with *DNMT3A*-CH at 14q32.13-*TCL1A*. The overall CH loci at 5p15.33-*TERT* (signals with lead variants rs2853677, rs13156167, and rs7705526), 3q25.33-*SMC4*, 6q21-*CD164*, and 11q22.3-*ATM* were also genome-wide significant for *DNMT3A*-mutant CH. We also found two novel loci for *DNMT3A*-CH marked by lead variants rs138994074 at 1q42.12-*PARP1* and rs8088824 at 18q12.3-*SETBP1* (Fig. 4b; Supplementary Table 18). The three *TET2-CH* associated loci included the lead variant rs2736100 at 5p15.33-*TERT*, that was moderately correlated (*r*^2^=0.44) with the overall CH lead variant rs2853677 in the same region. The other two risk loci, both new in the context of *TET2*-CH, were at lead variants rs10131341 (14q32.13-*TCL1A*) and rs79633204 (7q32.2-*TMEM209*; Fig. 4c; Supplementary Table 19). Notably, the A allele of rs10131341 had opposite associations with *TET2*-CH (OR=1.28, *P*=6.8×10^−10^) versus *DNMT3A*-CH (OR=0.87, *P*=6.4×10^−8^). A trend for opposite effects at 14q32.13-*TCL1A* was also observed in a previous study^17^, but did not achieve genome-wide significance for *TET2*-CH.

When comparing 4,049 individuals with large or 6,154 individuals with small clones against 173,918 controls of European ancestry without CH, we found that the overall CH loci at 5p15.33-*TERT* and 3q25.33-*SMC4* were associated at genome-wide significance with large clone CH (Fig. 4d; Supplementary Table 20), while 5p15.33-*TERT* and 6q21-*CD164* were associated with small clone CH. For small clone CH risk, we also identified a previously unreported locus marked by rs72755524 at 5p13.3 in a region with several lincRNAs (Fig. 4e; Supplementary Table 21). Additional signals suggested by approximate conditional analysis at each locus identified in this study are listed in Supplementary Table 17. Examining heterogeneity of associations across the five CH traits using forest plots (Extended Data Fig. 4) revealed that in addition to 14q32.13-*TCL1A*, the lead alleles at 6q21-*CD164* also had opposite effects on *DNMT3A*-versus *TET2*-CH. In addition, the lead variants at 6q21-*CD164* and 5p13.3-*LINC02064* were associated with small, but not large, clones while the association at 7q32.2-*TMEM209* was highly specific to *TET2*-CH.

Finally, in addition to our common variant GWAS, we performed a more focused scan to explore rare variant (MAF: 0.2%-1%) associations in each of three CH traits that included >5,000 European-ancestry individuals with CH (i.e., overall CH, *DNMT3A*-CH, and small clone CH; each compared to 173,918 controls) declaring associations significant at *P*<10^−9^. This identified one new locus at 22q12.1-*CHEK2* where the T allele (frequency=0.3%) of lead variant rs62237617 was perfectly correlated (*r*^2^=1) with the 1100delC CHEK2 protein-truncating allele (rs555607708) and conferred a large increase in risk of *DNMT3A* mutation-associated CH (OR=4.1, 95%CI: 2.7-6.1, *P*=6.3×10^−12^). The *CHEK2* c.1100delC frameshift mutation or its tagging variant rs62237617 are known to be associated with myeloproliferative neoplasms (MPNs) and *JAK2* V617F-driven CH (though not genome-wide significant for either trait)^18^, elevated white blood cell counts and plateletcrit^27^, as well as risk of prostate and breast cancers^28,29^. The *DNMT3A*-CH risk increasing alleles in the *CHEK2* and *PARP1* regions were also associated with later age at menopause in a recent analysis^30^, suggesting a role for inhibition of DNA damage sensing and apoptosis in both CH and reproductive ageing^31^.

### Genetic relationship between hematological chromosomal mosaicism and CH due to gene mutation

It is not known whether the germline genetic architecture underlying predisposition to CH due to individual gene mutations is similar to that underlying the risk of CH due to mosaic chromosomal alterations (mCAs). We used data from a recent GWAS of blood mCAs^32^ to answer this question and found that 13 of 19 unique lead variants identified for the five gene-mutant CH traits (overall, *DNMT3A, TET2*-, and large and small clone CH) were associated with hematological mCA risk at *P*<10^−4^ (Supplementary Table 22). Notably, for our lead variants rs2296312 (14q32.13-*TCL1A*) and rs8088824 (18q12.3-*SETBP1*), the alleles conferring increased *DNMT3A*-CH risk reduced hematological mCA risk (Supplementary Table 22). At the genome-wide level we found a correlation between overall CH and mCAs (*r*_g_=0.44, s.e.=0.21, *P*=0.037) using LDSC^22^. Further, a phenome-wide scan^33,34^ showed that several newly identified lead variants in our analyses were associated with multiple blood cell counts and traits (Supplementary Table 23).

### Gene-level associations and network analyses

We supplemented our GWAS with gene-level association tests for each of our five CH traits using two complementary methods: multi-marker analysis of genomic annotation (MAGMA) and a transcriptome-wide association study (TWAS) using blood-based *cis* gene expression quantitative trait locus data on 31,684 individuals^35^ and summary-based Mendelian randomization (SMR) coupled with the heterogeneity in dependent instruments colocalization test^36^. Both approaches converged on a new locus at 6p21.1, associated at gene-level genome-wide significance (*P*_MAGMA_<2.6×10^−6^, *P*_SMR_<3.2×10^−6^) with *DNMT3A*-mutant CH and marked by *CRIP3* (*P*_MAGMA_=3.4×10^−7^, *P*_SMR_=6.6×10^−7^; Fig. 3a; Supplementary Tables 24 and 25). While *CRIP3* is the only 6p21.1 gene to reach gene-level genome-wide significance in both MAGMA and SMR, we did find sub-threshold evidence for association between *SRF* or *ZNF318* in the same region and *DNMT3A*-mutant CH (Fig. 5a). Of note, *SRF* encodes the serum response factor that is known to regulate HSC adhesion^37^ while *ZNF318* is an occasional somatic driver gene for CH^38^. More globally, protein-protein interaction (PPI) network analysis^39^ using proteins encoded by the 57 genes with *P*_MAGMA_<0.001 in the overall CH analysis (Supplementary Table 24) as “seeds”, identified the largest sub-network (Fig. 5b) as encompassing 13/57 proteins with major hub nodes highlighted as TERT, PARP1, ATM, and SMC4. This was consistent with the emerging theme that key genes at sub-threshold GWAS loci for the same trait are often part of interconnected biological networks^40,41^. The sub-threshold genes identified by MAGMA that encoded protein hubs in this network included *FANCF* (DNA repair pathway) and *PTCH1* (hedgehog signaling; Fig. 5b), both implicated in the pathogenesis of acute myeloid leukemia^42,43^ and *GNAS*, a somatic driver of CH^44^. The CH sub-network (seeds and non-seed interacting proteins) was significantly enriched for several pathways of relevance to common disease including DNA repair, cell cycle regulation, telomere maintenance, and platelet homeostasis (Supplementary Table 26).

**Fig. 5:**
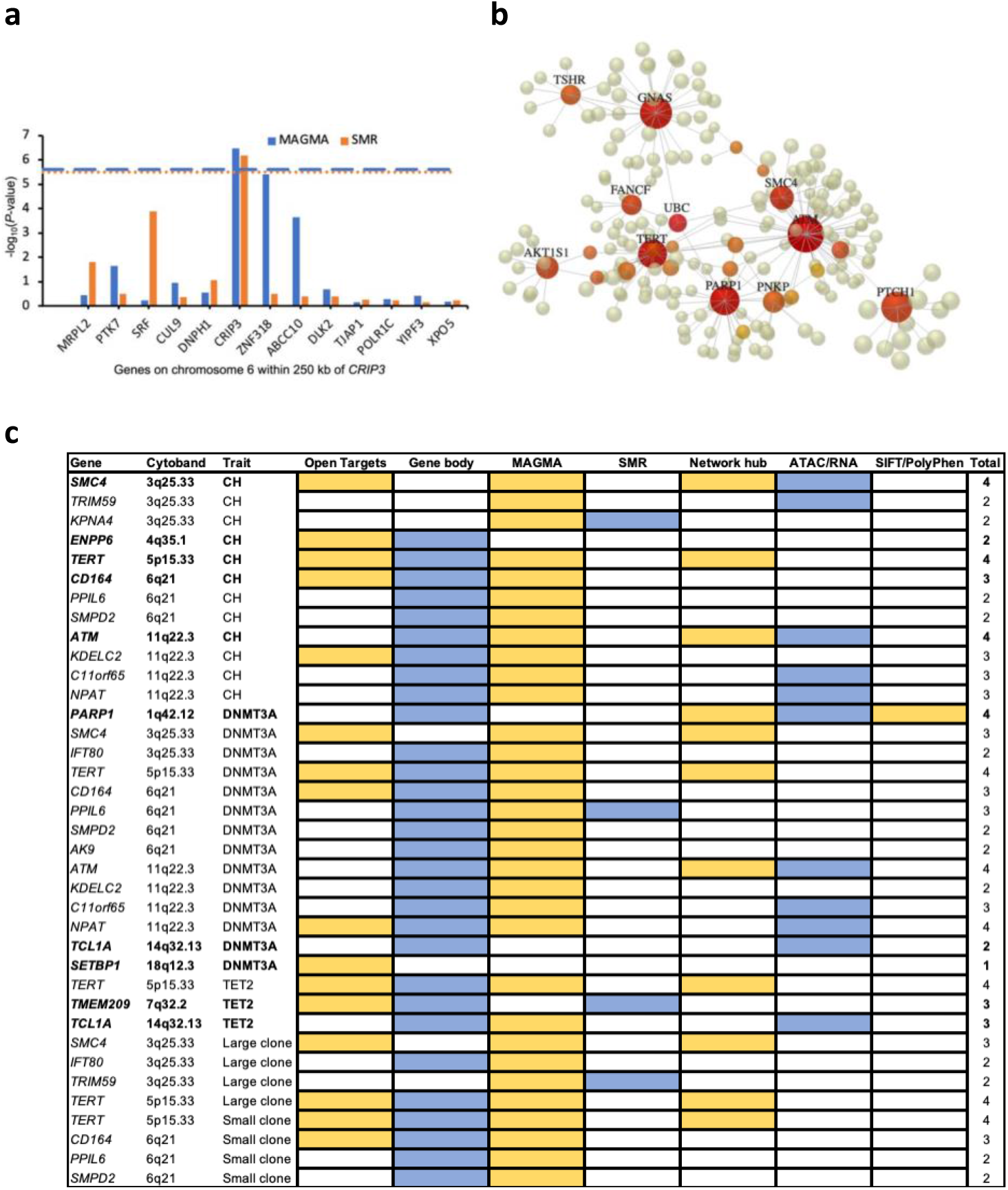
Gene-level association and protein-protein interaction network analyses, and functional target gene prioritization matrix. **a**, Gene-level associations in the 6q21 region within 250 kilobases of *CRIP3*, i.e., between GRCh37 positions 43,017,448 and 43,526,535 on chromosome 6. The X-axis lists all the genes in this region that were tested by both MAGMA and SMR. *CRIP3* was the only gene located more than one megabase away from a GWAS-identified lead variant that was found to be associated with CH at gene-level genome-wide significance by both MAGMA and SMR. The Y-axis depicts the *P*-value (-log_10_) for association in the MAGMA and SMR analyses. The gene-level genome-wide significance threshold in MAGMA (*P*=2.6×10^−6^ after accounting for 19,064 genes tested) is indicated by the blue dashed line and in SMR (*P*=3.2×10^−6^ after accounting for 15,672 genes tested) by the orange dotted line. Both *CRIP3* and *SRF* had SMR HEIDI *P*>0.05 indicating colocalization of the GWAS and expression quantitative trait locus associations. **b**, Largest sub-network of genes/proteins associated with overall CH risk. All genes (n=57) with *P*_MAGMA_<0.001 in the overall CH MAGMA analysis were mapped to proteins and used as “seeds” for network construction that was done by integrating high-confidence protein-protein interactions from the STRING database. The largest sub-network constructed contained 13 of the 57 seed proteins and included 210 nodes and 231 edges. The colored nodes indicate seed proteins that interact with at least two other proteins in this sub-network with the intensity of redness increasing with number of interacting proteins. Seed proteins that interact with six or more other proteins in the sub-network are named above their corresponding node. c, Matrix of target genes (protein coding) prioritized by seven approaches (Open Targets, fine-mapped variant-gene body overlap, MAGMA, SMR, network analysis hub, fine-mapped variant-ATAC-seq peak overlap followed by ATAC-RNA-seq correlation, and SIFT/PolyPhen scores) across the loci identified in this study. Only genes prioritized by at least two methods are shown, with the exception of *SETBP1* that is shown despite being prioritized by only one method since it was the only gene prioritized at 18q12.3 and also happens to be an occasional somatic driver of CH. There were no protein coding genes within 1 Mb of the new small clone CH lead variant rs72755524 at 5p13.3 (nearest non-protein coding gene: *LINC02064*, nearest protein coding gene: *CDH6*) and this locus is not shown in the matrix. Abbreviations: MAGMA, multi-marker analysis of genomic annotation; SMR, summary-based Mendelian randomization; HEIDI, heterogeneity in dependent instruments test; SIFT, Sorting Tolerant From Intolerant.

### Functional target gene prioritization at CH risk loci

In order to prioritize putative functional target genes at the *P*_lead-variant_<5×10^−8^ loci identified by our GWAS of five CH traits, we combined gene-level genome-wide significant results from MAGMA and SMR (Supplementary Tables 24 and 25) with five other lines of evidence: PPI network hub status of the gene (Supplementary Table 27), variant-to-gene searches of the Open Targets database^45^ for lead variants, overlap between fine-mapped variants^46,47^ (Supplementary Table 28) and (i) gene bodies, (ii) regions with accessible chromatin correlated with nearby gene expression in hematopoietic progenitor cells^24,48–50^, and (iii) missense variant annotations^51,52^ (Supplementary Table 29). Genes nominated by at least two of these approaches are listed in Fig. 5c. The genes nominated by the largest number of approaches, and representing the most likely targets, were *SMC4, ENPP6, TERT, CD164, ATM, PARP1, TCL1A, SETBP1*, and *TMEM209*.

Among the newly identified loci, *CD164* codes for Sialomucin core protein 24, a cell adhesion molecule that regulates HSC adhesion, proliferation, and migration^53,54^. Lead variant rs138994074 at 1q42.12 was strongly correlated (*r*^2^=0.93) with rs1136410, a missense germline mutation in *PARP1* (Supplementary Table 29) wherein the G allele, which is protective for *DNMT3A*-CH, leads to a missense variant (p.Val762Ala) in the catalytic domain of its protein product associated with reduced Poly (ADP-ribose) polymerase-1 activity^55^. While *SETBP1* was only nominated by one approach (Open Targets^45^) and was the only gene nominated at 18q12.3, its nomination is strengthened by the fact that somatic *SETBP1* mutations are recognized drivers of myeloid malignancies^56,57^.

### Mendelian randomization (MR) to uncover the causes and consequences of CH

We integrated several large GWAS datasets (Supplementary Tables 30 and 31) and used two-sample inverse-variance-weighted MR^58^ to appraise putative causes and consequences of CH. Genetically-predicted smoking initiation^59^ was associated with overall CH risk (OR=1.15, 95%CI: 1.05-1.25, *P*=2.2×10^−3^). Point estimates of the effect size were consistent in direction across MR analyses for *DNMT3A, TET2*, and large and small clone CH (Fig. 6a; Supplementary Table 32), with the largest odds ratio observed for large clone CH (OR=1.24). We also appraised the roles of leukocyte telomere length (LTL)^60^, alcohol use^59^, adiposity^61^, genetic liability to T2D^62^, major circulating lipids^63^, blood-based epigenetic aging phenotypes^64^, blood cell counts and indices^27^, and circulating cytokines and growth factors^65^ as potential risk factors for CH using MR (Fig. 6; Supplementary Tables 32, 33, and 34 for full results, including sensitivity analyses). Genetically predicted longer LTL was associated with increased overall CH risk (OR=1.56, 95%CI: 1.25-1.93, *P*=5.7×10^−5^), an association that was also seen with *DNMT3A*-, *TET2*-, and large and small clone CH (Fig. 6b; Supplementary Table 32). We found that higher genetically predicted BMI was associated with increased risk of large clone CH (OR=1.15, 95%CI: 1.01-1.31, *P*=0.029). Genetically elevated circulating apolipoprotein B levels were associated with increased (OR=1.18, 95%CI: 1.01-1.36, *P*=0.032; Fig. 6c), whilst genetically predicted alcohol use was associated with decreased (OR=0.46, 95%CI: 0.25-0.83, *P*=0.010) risk of *TET2*-CH. Among cytokines, genetically-elevated circulating macrophage inflammatory protein 1a, a regulator of myeloid differentiation and HSC numbers^66^, was associated with risk of *DNMT3A*-CH (OR=1.13, 95%CI 1.03-1.23, *P*=7.1×10^−3^; Supplementary Table 34).

**Fig. 6:**
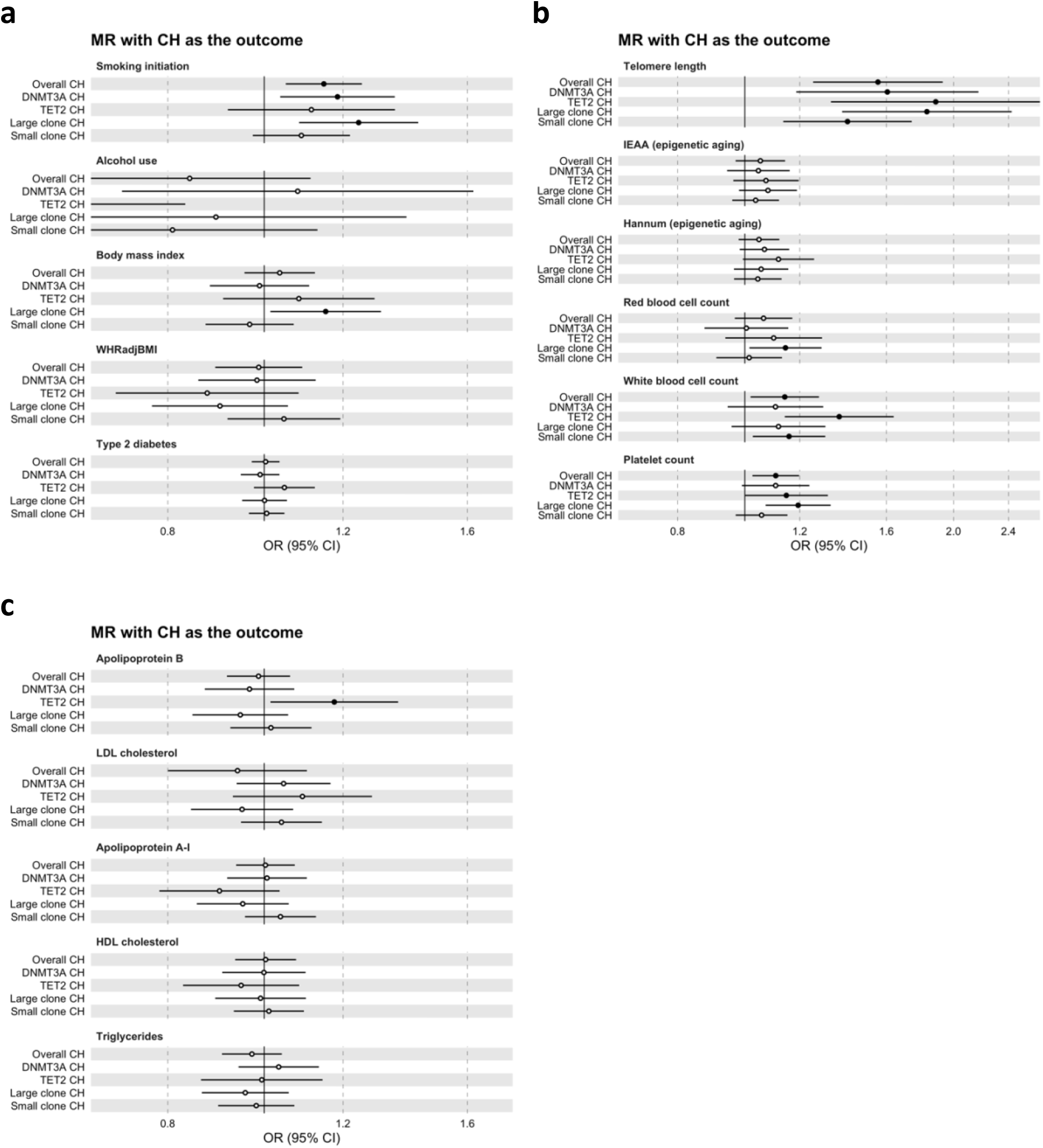
Two-sample inverse-variance-weighted Mendelian randomization forest plots with CH traits as outcomes. Odds ratios (ORs) for CH risk are represented as per (i) standard deviation unit for continuous exposures (alcohol use in drinks per week, body mass index, waist-to-hip ratio adjusted for body mass index (WHRadjBMI) **(a);** leukocyte telomere length, two epigenetic aging traits, red cell, white cell and platelet counts **(b)**, and five circulating lipid traits **(c))** and (ii) log-odds unit for binary exposures (smoking initiation (ever having smoked regularly) and genetic liability to type 2 diabetes **(a))**. Details of units are provided in Supplementary Table 30. OR markers with corresponding *P*-value<0.05 are represented by filled circles. Error bars represent 95% confidence intervals (CIs). Full results, including sensitivity analyses, are presented in Supplementary Tables 32, 33, and 34. Abbreviations: MR, Mendelian randomization; CH, clonal hematopoiesis; WHRadjBMI, waist-to-hip ratio adjusted for body mass index; LDL, low-density lipoprotein cholesterol; HDL, high-density lipoprotein cholesterol; IEAA, intrinsic epigenetic age acceleration.

We used independent (*r*^2^ < 0.001) variants associated with overall, *DNMT3A, TET2*, and large and small clone CH at *P*<10^−5^ as genetic instruments for each of these traits and assessed their associations with outcomes (Supplementary Tables 31, 35, and 36 for full results, including sensitivity analyses). Since more variants were available at *P*<5×10^−8^ for overall and for *DNMT3A* CH, we also examined the consistency of associations when using genome-wide (GWS; *P*<5×10^−8^) and sub-genome-wide significant (sub-GWS; *P*<10^−5^) instruments for these two traits. Using the sub-GWS instrument, genetic liability to overall CH had the largest associations (Fig. 7a) with MPN risk^48^ (OR=1.99, 95%CI: 1.23-3.23, *P*=5.4×10^−3^), intrinsic epigenetic age acceleration^64^ (IEAA, which represents a core characteristic of HSCs^67^; beta= 0.39, 95%CI: 0.08-0.69, *P*=0.01) and the blood-based Hannum epigenetic clock^64^ (beta= 0.27, 95%CI: 0.04-0.49, *P*=0.02). Larger associations were observed when using the GWS instrument (Fig. 7a) and the direction of these was consistent when evaluating genetic liability to *DNMT3A, TET2*, and large and small clone CH as exposures (Supplementary Tables 35 and 36). Genetic liability to CH conferred increased risks of lung^68^, prostate^69^, ovarian^70^, oral cavity/pharyngeal^71^, and endometrial cancers^72^ (Fig. 7; Supplementary Table 35) with the strongest associations observed between overall CH and lung (OR=1.17, 95%CI: 1.05-1.29, *P*=2.9×10^−3^); *DNMT3A*-CH and prostate (OR=1.08, 95%CI:

**Fig. 7:**
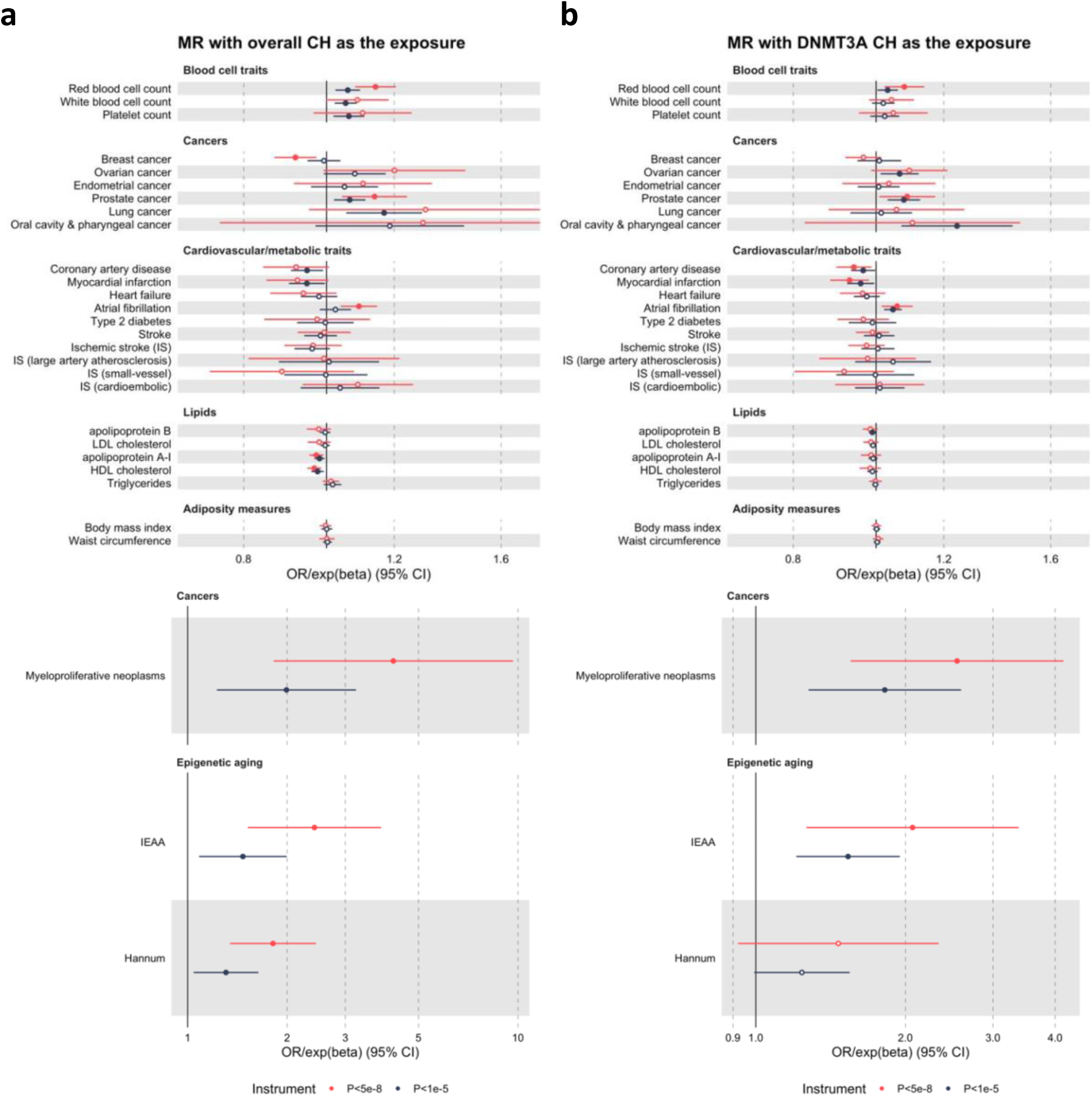
Two-sample inverse-variance-weighted Mendelian randomization forest plots with CH traits as exposures. Forest plots with odds ratio (OR) markers (for cancers and cardiovascular/metabolic traits) or exponentiated beta coefficient (exp(beta)) markers (for blood cell traits, lipids, adiposity measures, and epigenetic aging indices). ORs/exp(betas) are represented as per log-odds unit increase in genetic liability to **a**, overall CH, or **b**, DNMT3A CH. OR/exp(beta) markers with corresponding *P*-value<0.05 are represented by filled circles. Error bars represent 95% confidence intervals (CIs). Red markers and error bars represent results using genetic instruments comprised exclusively of genome-wide significant (*P*<5×10^−8^) variants. Black markers and error bars represent results when using genome-wide significant and sub-genome-wide significant (*P*<10^−5^) variants in the genetic instrument. Large effect size estimates (ORs/exp(betas)) are shown in the lower panels. Full results, including sensitivity analyses, are presented in Supplementary Tables 35 and 36. Abbreviations: MR, Mendelian randomization; IS, ischemic stroke; LDL, low-density lipoprotein cholesterol; HDL, high-density lipoprotein cholesterol; IEAA, intrinsic epigenetic age acceleration.

1.03-1.13, *P*=8.6×10^−4^), ovarian (OR=1.07, 95%CI: 1.01-1.12, *P*=0.015), and oral cavity/pharyngeal (OR=1.24, 95%CI: 1.07-1.44, *P*=4.4×10^−3^); and *TET2*-CH and endometrial (OR=1.05, 95%CI: 1.00-1.09, *P*=0.033) cancers. MR analyses did not support causal risk-conferring associations between genetic liability to CH and CAD^73^, ischemic stroke^74^, and heart failure^75^ with similar lack of evidence across gene-specific and clone size-specific CH, and GWS instrument analyses (Fig. 7; Supplementary Table 35). However, we did uncover an association between genetic liability to overall CH or *DNMT3A*-CH and atrial fibrillation^76^ risk (OR=1.09, 95%CI: 1.04-1.15, *P*=4.9×10^−4^ for overall CH with the GWS instrument; Supplementary Table 35). Among cytokines and growth factors^65^, genetic liability to overall CH was associated with elevated circulating stem cell growth factor beta (beta= 0.19; 95%CI: 0.07-0.30, *P*=1.1×10^−3^). MR analyses also revealed bidirectional associations between CH phenotypes and several blood cell counts and traits^27^, suggesting a shared underlying genetic liability to CH and pan-blood cell proliferation (Figs. 6b and 7; Supplementary Tables 33 and 35). Finally, we found little evidence to support an association between genetic liability to CH and LTL (Supplementary Table 36), indicating that longer LTL was a cause rather than a consequence of CH.

## Discussion

We present a large observational and genetic epidemiological analysis of CH and report a series of novel insights into the causes and consequences of this common aging-associated phenomenon. We increase the number of germline associations with CH in European-ancestry populations from four^17^ to 14, reveal heterogeneity of associations by CH driver gene and clone size, and implicate putative new CH susceptibility genes, including *CD164, ATM* and *SETBP1*, through functional annotation. We also demonstrate that the CH GWAS signal is enriched at epigenetic marks specific to the hematopoietic system. The robustness of our GWAS analysis is further affirmed by our replication of previous European ancestry-specific CH associations^17^, the consistency of our estimates of CH heritability with previous reports^17,77^, and the fact that many of our lead variants are associated with related traits^27,32,60,78^.

New CH risk loci included the *PARP1* coding variant rs1136410, where the G allele is protective for *DNMT3A*-CH and associated with reduced catalytic activity^55^ suggesting that this most common form of CH may be vulnerable to PARP inhibition, in keeping with the observed synergy between PARP and DNMT inhibitors^79^. At 14q32.13-*TCL1A*, we replicate the reported association with *DNMT3A*-CH^17^ and identify a new genome-wide significant association with *TET2*-CH. Strikingly, however, we found that the association operates in the opposite direction for *TET2-CH*, versus *DNMT3A*-CH. This inverse relationship is tantalizing in light of recent observations that ageing has different effects on the dynamics of these two forms of CH, resulting in *TET2* CH becoming more prevalent than *DNMT3A* CH in those aged over 80 years^80,81^. Also notable in this light, is the finding of an association at the *CD164* locus with *DNMT3A*, and a trend in the opposite direction for *TET2*-CH. As *CD164* is expressed in the earliest HSCs^53^ and encodes an important regulator of HSC adhesion^54,82^, this proposes that HSC migration and homing may play important roles in CH pathogenesis. The reciprocal relationship of both *TCL1A* and *CD164* with the two main CH subtypes, suggests that their expression needs to be tightly regulated to prevent the development of one or other subtype of CH, making these loci important targets for hijack by the effects of somatic mutations.

The rich phenotypic data captured by the UKB, coupled with our genetic analysis of CH and external GWAS datasets, enabled us to explore associations of CH using multivariable regression and interrogate, at scale, potential causal relationships between CH and its putative risk factors and consequences using MR. This highlighted for the first time that smoking and longer telomere length are causal risk factors for CH. These associations were valid across multiple CH subtypes and, in the case of smoking, corroborated by observational estimates. We also reveal that not only is genetic predisposition to CH causally associated with MPN risk, but it also increases the risk of lung, prostate, ovarian, oral/pharyngeal, and endometrial cancers. In these analyses, the use of two-sample MR protected against potential reverse causality arising from cancer therapy-induced selection pressure on hematopoietic clones^83^. These MR results suggest that genetic liability to CH may be a biomarker for development of cancer elsewhere in the body. An analogous relationship has previously been identified by MR for the association of genetic predisposition to Y chromosome loss in blood and solid tumor risk^31^.

We investigated the recently identified association of CH with blood-based epigenetic clocks^84^, using bi-directional MR and show that this association is likely to be causal in the direction from CH to epigenetic age acceleration. We also showed that genetic predisposition to CH was associated with elevated circulating levels of stem cell growth factor beta, a secreted sulfated glycoprotein that regulates primitive hematopoietic progenitor cells^85^. Finally, we unraveled a previously unreported association between genetic liability to CH and atrial fibrillation risk, which was also supported by our observational analysis. However, unlike previous reports based on significantly smaller sample numbers^5,10,21^, we did not find evidence in observational and MR analyses to support an association between CH and CAD or ischemic stroke risk. However, our MR analyses indicated that higher BMI and circulating apolipoprotein B levels were associated with *TET2* and large clone CH risks, respectively, with apolipoprotein B being the key causal lipid risk factor for CAD^63,86^. These associations taken together with the fact that age and smoking are strong risk factors for CH raise the possibility that previously reported associations of CH with CAD and stroke risks may suffer from residual confounding.

Collectively, our findings substantially illuminate the landscape of inherited susceptibility to CH and provide new insights into the causes and consequences of CH with implications for human health and ageing.

## Methods

### Study population and exome sequence data

The United Kingdom Biobank (UKB) is a prospective longitudinal study containing in-depth genetic and health information from half a million UK participants. For this study, we have selected 200,453 individuals (200k) who had whole exome sequencing (WES) data available (age range: 38-72, median age: 58; 55% female; 83% White British). WES was generated in two batches, the first of approximately 50,000 samples (50k)^87^ and the second comprising an additional 150,000 samples (150k)^19^. Exomes were captured using the IDT xGen Exome Research Panel v1.0 including supplemental probes; a different IDT v1.0 oligo lot was used for each batch. Multiplexed samples were sequenced with dual-indexed 75×75 bp paired-end reads on the Illumina NovaSeq 6000 platform using S2 (50k samples) and S4 (150k samples) flow cells. The 50k samples were firstly computed using FE protocol and reprocessed later to match the second batch of 150k sequences that were processed using a new improved unified OQFE pipeline. As the initial 50k samples were sequenced on S2 flow cells and with a different IDT v1.0 oligo lot than the remaining 150k samples, which were sequenced on S4 flow cells, we included the WES batch as a covariate in downstream analyses.

The UK Biobank study has been approved by the North West Multicentre Research Ethics Committee (11/NW/0382). All participants provided written informed consent. The current study has been conducted under approved UK Biobank application numbers 56844 and 29202.

### Whole exome sequence data processing, CH mutation calling and filtering

CRAM files generated by the OQFE pipeline were obtained from UKB (Fields 23143-23144; www.ukbiobank.ac.uk). Variant-calling on WES data from 200,453 individuals was performed using Mutect2, Genome Analysis Toolkit (GATK) version 4.1.8.1^88^. Briefly, Mutect2 was run in “tumor-only” mode with default parameters, over the exon intervals of 43 genes previously associated with CH (Supplementary Table 1). To filter out potential germline variants we used a population reference of germline variants generated from 1000 Genomes Project (1000GP)^89^ and the Genome Aggregation Database (gnomAD)^90^. All resources were obtained from the GATK Best practices repository (gs://gatk-best-practices/somatic-hg38). Raw variants called by Mutect2 were filtered out with *FilterMutectCalls* using the estimated prior probability of a reading orientation artefact generated by *LearnReadOrientationModel* (GATK v.4.1.8.1). Putative variants flagged as ‘PASS’ using FilterMutectCalls or flagged as ‘germline’ if present at least 2 times with the ‘PASS’ flag in other samples were selected for filtering. Gene annotation was performed using Ensembl Variant Effect Predictor (VEP) (v.102)^91^. We required variants with a minimum number of alternate reads of 2, evidence of the variant on both forward and reverse strand, a minimum depth of 7 reads for SNVs and 10 reads for short indels and substitutions and a minor allele frequency (MAF) lower than 0.001 (according to 1000GP phase 3 and gnomAD r2.1). For new variants, not previously described in the Catalogue of Somatic Mutations in Cancer (COSMIC; v.91)^92^ nor in the Database of Single Nucleotide Polymorphisms (dbSNP; build 153)^93^, we used a minimum allele count per variant of 4, and a MAF lower than 5×10^−5^. From resulting variants, we selected those that: i) are included in a list of recurring hotspots mutations associated with CH and myeloid cancer (Supplementary Table 2); ii) have been reported as somatic mutations in hematological cancers at least 7 times in COSMIC; or iii) met the inclusion criteria of a predefined list of putative CH variants, previously described^17,77^ (Supplementary Table 3). We included previous variants flagged as germline by *FilterMutectCalls* if: 1) the number of cases in the cohort flagged as germline were lower than the ones flagged as PASS; and 2) at least one of the cases had a *P*<0.001 for a one-sided exact binomial test, where the null hypothesis was that the number of alternative reads supporting the mutation were 50% of the total number of reads (95% for copy number equal to one), except for hotspot mutations that were all included. For the final list, we excluded all variants not present in COSMIC nor in the list of hotspots that had a MAF equal or higher than 5×10^−5^ and either the mean variant allele fraction (VAF) of all cases was higher than 0.2 or the maximum VAF was lower than 0.1. Frameshift, nonsense, and splice-site mutations not present in COSMIC nor in the hotspot list were further excluded if for each variant none of the cases had a *P*<0.001 for a one-sided exact binomial test. A complete list of filtered variants is provided in Supplementary Table 4.

### Trait selection and modelling for the conventional observational multivariable regression analyses

Phenotypes were downloaded in December 2020 and individual traits were pulled out from the whole phenotype file. Cancer, metabolic and cardiovascular disease (CVD) traits were generated combining individual traits and diagnosis dates based on disease definitions (Supplementary Table 8). For each definition of disease, the first diagnosis event that occurred in each trait was selected. Baseline was defined as the date of sample collection. The prevalent cases are those identified before the baseline, while incidence was defined as the events that occurred after the baseline. Unless specified, all regression models included age, sex, smoking status, WES batch and the first ten ancestry principal components as covariates. Blood cell counts and biochemical traits were log_10_ transformed and analyzed using a linear regression model, including the assessment center as covariate and, in the case of cholesterol and cholesterol species, the use of cholesterol lowering medication. Individuals with myeloid malignancies or hematological neoplasms at baseline were excluded from the analysis. For cancer, CVD and death risk, we performed a time-to-event regression analysis using the Cox proportional hazards model. The cancer/CVD/death event was used as an outcome and CH was considered as the exposure in these analyses. For CVD and death risk analyses, we also included body mass index, high-density lipoprotein cholesterol, low-density lipoprotein cholesterol, triglycerides, type 2 diabetes status, and hypertension status as covariates. Individuals with myeloid or other malignant neoplasms at baseline were excluded from all previous analyses. For associations between International Statistical Classification of Diseases and Related Health Problems 10th Revision (ICD-10) codes and CH status, a logistic regression model was used including age, sex, WES batch and the first ten ancestry principal components as covariates. Analyses were performed over the selected ICD-10 codes corresponding to diseases conditions (A to N), symptoms, signs, and abnormal clinical and laboratory findings (R) and factors influencing health status (Z). All analyses were performed using glm (R stats package v.4.0.2) and coxph (R survival package v.3.2-11) functions.

### Germline genotype data processing and genome-wide association analyses

Germline genotype data used were from the UKB release that contained the full set of variants imputed into the Haplotype Reference Consortium^94^ and 1000GP^89^ reference panels and genotyped on the UK BiLEVE Axiom Array or UKB Axiom Array^95^. Derivation of the analytic sample for UK Biobank of individuals of European ancestries followed quality control (QC) steps described previously^27^: after filtering genetic variants (call rate≥99%, imputation quality info score>0.9, Hardy-Weinberg equilibrium *P*-value≥10^−5^) and participants (removal of genetic sex mismatches), we excluded participants having non-European ancestries (self-report or inferred by genetics) or excess heterozygosity (>3 standard deviations from the mean), and included only one of each set of related participants (third-degree relatives or closer). After QC, we were left with 10,203 individuals with CH and 173,918 individuals without CH. The subset with CH included 5,185 and 2,041 individuals with *DNMT3A* and *TET2*-mutant CH, respectively, and 4,049 and 6,154 individuals with large (VAF≥0.1) and small (VAF<0.1) clone size CH, respectively. Association analyses were performed using non-infinitesimal linear mixed models implemented in BOLT-LMM^96^ with age at baseline, sex, and first 10 genetic principal components included as covariates.

Statistically independent lead variants for each CH phenotype were defined using linkage disequilibrium (LD)-based clumping with an *r*^2^ threshold of 0.05 applied across all genotyped and imputed variants with *P*<5×10^−8^, imputation quality score>0.6, and MAF>1%. This was implemented using the FUMA pipeline^97^. For the rare variant association scan, we used more stringent cut-offs of *P*<10^−9^ and imputation quality score>0.8 to define lead variants but did not require LD-clumping since only one such association was identified. Approximate conditional analysis conditioning on the common (MAF>1%) lead variants was performed using the *--cojo-cond* flag in the Genome-wide Complex Trait Analysis (GCTA) v1.93 tool^25,98^.

### Linkage disequilibrium score regression (LDSC)

We used LDSC^22^ to estimate the narrow-sense heritability of CH on the liability scale assuming the population prevalence of CH to be 10% (based on the prevalence of CH in the UKB “200k” cohort as shown in Fig. 1b) and constraining the LDSC intercept to 1. The intercept, which in its unconstrained form protects from bias due to population stratification, was constrained to 1 to provide more precise estimates given that there was little evidence of inflation in test statistics due to population structure in unconstrained analysis (unconstrained intercept estimated as 1.009 (s.e.=0.0067) and lambda genomic control factor of 0.999). We used the pre-computed 1000 Genomes phase 3 European ancestry reference panel LD score data set downloaded from alkesgroup.broadinstitute.org/LDSCORE for heritability estimation. We used the same LD scores and the *--rg* flag in LDSC to estimate the genetic correlation between the CH and mosaic chromosomal alteration GWAS summary statistics^32^. Cell-type group partitioned heritability analysis was performed as described in github.com/bulik/ldsc/wiki/Partitioned-Heritability using LD scores partitioned across 220 cell-type-specific annotations that were divided into 10 groups as previously described^23^: central nervous system, cardiovascular, kidney, adrenal/pancreas, gastrointestinal, connective/bone, immune/hematopoietic, skeletal muscle, liver, and other. Each of the 10 groups contained cell-type-specific annotations for four histone marks: H3K9ac, H3K27ac, H3K4me1, and H3K4me3^23^. We also used LD scores annotated as previously described^99^ based on open chromatin state (Assay for Transposase-Accessible Chromatin (ATAC)-seq) profiling by Corces et al.^24^ in various hematopoietic progenitor cells and lineages at different stages of differentiation.

### Gene-based and transcriptome-wide association studies, and network analyses

We undertook genome-wide gene-level association analyses using two complementary approaches. First, we used multi-marker analysis of genomic annotation (MAGMA) that involves mapping germline variants to the genes they overlap, accounting for LD between variants, and performing a statistical multi-marker association test^100^. Second, we performed a transcriptome-wide association study (TWAS) using blood-based *cis* gene expression quantitative trait locus (eQTL) data on 31,684 individuals^35^ and summary-based Mendelian randomization (SMR) coupled with the heterogeneity in dependent instruments (HEIDI) colocalization test to identify germline genetic associations with CH risk mediated via the transcriptome^36^. The gene-level genome-wide significance threshold in the MAGMA analyses was set at *P*=2.6×10^−6^ to account for testing 19,064 genes and for SMR was set at *P*=3.2×10^−6^ after adjustment for testing 15,672 genes. Further, only genes with SMR *P*<3.2×10^−6^ and HEIDI *P*>0.05 were declared genome-wide significant in the SMR analyses since the HEIDI *P*>0.05 strongly suggests colocalization of the GWAS and eQTL signals for a given gene^36^. The NetworkAnalyst 3.0^39^ webtool available at www.networkanalyst.ca was used for network analysis. All genes with *P*<10^−3^ in each MAGMA analysis for overall, *DNMT3A* and *TET2*-mutant, and large and small clone CH were used as input. The protein-protein interactome selected was STRING v10^101^ with the recommended parameters (confidence score cut-off of 900 and requirement for experimental evidence to support the protein-protein interaction). The largest possible network was constructed from the seed genes/proteins and the interactome proteins as previously described^39^. Hub nodes were defined as nodes with degree centrality≥10 (i.e., nodes with at least 10 edges or connections to other proteins in the network as a measure of its importance in the network and consequently its biology). Pathway analysis of this largest network was conducted using the enrichment tool built into the NetworkAnalyst webtool and with the Reactome pathway repository^102^.

### Fine-mapping and target gene prioritization

We fine-mapped the lead variant signals identified by the FUMA LD-clumping pipeline using the Probabilistic Identification of Causal Single Nucleotide Polymorphisms (PICS2) algorithm^46,47^ to identify candidate causal variants most likely to underpin each association. The PICS2 algorithm and webtool (pics2.ucsf.edu) computes the likelihood that each variant in LD with the lead variant is the true causal variant in the region by leveraging the fact that for variants associated merely due to LD, the strength of association scales asymptotically with correlation to the true causal variant^46^. We only retained variants with a PICS2 probability of 1% or more in our final list of fine-mapped candidate causal variants. We overlapped these fine-mapped variants with gene body annotations as previously described^48^ using GENCODE release 33^103^ (build 37) annotations after removing ribosomal protein genes (code and data adapted from github.com/sankaranlab/mpn-gwas). Fine-mapped variants were also overlapped with ATAC-seq peaks across 16 hematopoietic progenitor cell populations and ATAC-RNA count correlations calculated using Pearson coefficients for hematopoietic progenitor cell RNA counts of genes within 1 Mb of the ATAC peaks were used to identify putative target genes of fine-mapped variants that overlapped ATAC-seq peaks. This pipeline has been used and described extensively before^24,48–50^, and we adapted the code and data for the pipeline from github.com/sankaranlab/mpn-gwas. We also looked up the SIFT^51^ and PolyPhen^52^ scores for these fine-mapped variants using the SNPnexus v4 web-based annotation tool (www.snp-nexus.org/v4)^104^ to identify coding variants with predicted functional consequences. Finally, we used the Open Targets Genetics resource^45^ (genetics.opentargets.org) to identify the most likely target gene of the lead variant at each locus as per Open Targets and used this in our omnibus target gene prioritization scheme described below.

In order to prioritize putative target genes at the *P*_lead-variant_<5×10^−8^ loci identified by our GWAS of overall CH, *DNTM3A*-CH, *TET2*-CH and large/small clone size CH, we combined gene-level genome-wide significant results from (1) MAGMA and (2) SMR with (3) protein-protein interaction network hub status of the gene, (4) variant-to-gene searches of the Open Targets database for lead variants, and overlap between fine-mapped variants and (5) gene bodies, (6) regions with accessible chromatin (ATAC-seq peaks) across 16 hematopoietic progenitor cell populations that were also correlated with nearby gene expression (RNA-seq) in the same cell populations, and (7) missense variant annotations from SIFT and PolyPhen. Genes nominated by at least two of the seven approaches were listed (except where only one of the seven methods nominated a single gene in a region in which case that gene was listed) and the genes nominated by the largest number of approaches represented the most likely targets at each locus.

### Phenome-wide association scan for lead variants

We used PhenoScanner V2^33,34^ available at www.phenoscanner.medschl.cam.ac.uk with catalogue set to “diseases & traits”, p-value set to “5E-8”, proxies set to “EUR” and *r*^2^ set to “0.8” to search for published phenome-wide associations between our lead variants or variants in strong linkage disequilibrium (*r*^2^>0.8) with the lead variants and other diseases and traits.

### Mendelian randomization analysis

Mendelian randomization (MR)^105,106^ uses germline variants as instrumental variables to proxy an exposure or potential risk factor and evaluate evidence for a causal effect of the exposure or potential risk factor on an outcome. Due to the random segregation and independent assortment of alleles at meiosis, MR estimates are less susceptible to bias from confounding factors as compared to conventional observational epidemiological studies. As the germline genome cannot be influenced by the environment after conception or by preclinical disease, MR estimates are also less susceptible to bias due to reverse causation. MR estimates represent the association between genetically predicted levels of exposures or risk factors and outcomes, as compared to conventional observational epidemiological estimates, which represent direct associations of the exposure or risk factor levels with outcomes. Effect allele harmonization across GWAS summary statistics datasets followed by two-sample Mendelian randomization analyses were performed using the TwoSampleMR v0.5.6 R package^58^. The CH phenotypes were considered as both exposures (to identify consequences of genetic liability to CH) and outcomes (to identify risk factors for CH). When considering CH phenotypes as outcomes, germline variants associated with putative risk factors or exposures at *P*<5×10^−8^ were used as genetic instruments for the risk factors/exposures, except for the appraisal of circulating cytokines and growth factors^65^ wherein variants associated with cytokines/growth factors at *P*<10^−5^ were used as instruments. Inverse-variance weighted analysis^107^ was the primary analytic approach with pleiotropy-robust sensitivity analyses carried out using the MR-Egger^108^ and weighted median^109^ methods. A full list of external GWAS data sources used for MR analyses is provided in Supplementary Tables 30 and 31.

## Data Availability

Individual-level UK Biobank data used in this study can be requested via application to the UK Biobank (https://www.ukbiobank.ac.uk). Publicly-available genome-wide association study summary statistics used in this study can be accessed via the IEU OpenGWAS project (gwas.mrcieu.ac.uk).

## Acknowledgements

This work was funded by a joint grant from the Leukemia and Lymphoma Society (RTF6006-19) and the Rising Tide Foundation for Clinical Cancer Research (CCR-18-500), and by the Wellcome Trust (WT098051). SPK is supported by a United Kingdom Research and Innovation (UKRI) Future Leaders Fellowship (MR/T043202/1) and leads the somatic genomics theme of the Integrative Cancer Epidemiology Programme at the University of Bristol that is funded by Cancer Research UK (C18281/A29019). PMQ is funded by the Miguel Servet Program (CP20/00130). MAF is funded by a Wellcome Clinical Research Fellowship (WT098051). RL is supported by Cancer Research UK (C18281/A29019). PC is supported by a British Heart Foundation Clinical Training Research Fellowship. SB is supported by a Sir Henry Dale Fellowship jointly funded by the Wellcome Trust and the Royal Society (204623/Z/16/Z). GSV is supported by a Cancer Research UK Senior Cancer Fellowship (C22324/A23015) and work in his lab is also funded by the European Research Council, Kay Kendall Leukaemia Fund, Blood Cancer UK, and the Wellcome Trust. This research was conducted using the UK Biobank resource under applications 56844 and 29202. We thank the participants and investigators involved in the UK Biobank resource and in the other genome-wide association studies cited in this work who collectively made this research possible.

## Author Contributions

SPK, PMQ, and GSV conceived, designed, and supervised the study. SPK and PMQ carried out data analyses and generated tables and figures. MG, MSV, and MAF helped with mutation calling and filtering. TJ and SB performed genome-wide association analyses. RL assisted with Mendelian randomization analyses. VI helped with UK Biobank data access and handling. SB and PC advised on Mendelian randomization analyses. CB and PC helped with UK Biobank trait selection and filtering. SPK, PMQ, and GSV drafted the manuscript with inputs from all authors. All authors approved the final version of the paper.

## Data Availability

Individual-level UK Biobank data can be requested via application to the UK Biobank (https://www.ukbiobank.ac.uk). The CH call set will be returned to the UK Biobank to enable individual-level data linkage for approved UK Biobank applications.

## Code Availability

Code used in this study is available at https://github.com/pmquiros/CH_UKBiobank and https://github.com/siddhartha-kar/clonal-hematopoiesis.

---

**Extended Data Fig. 1:**
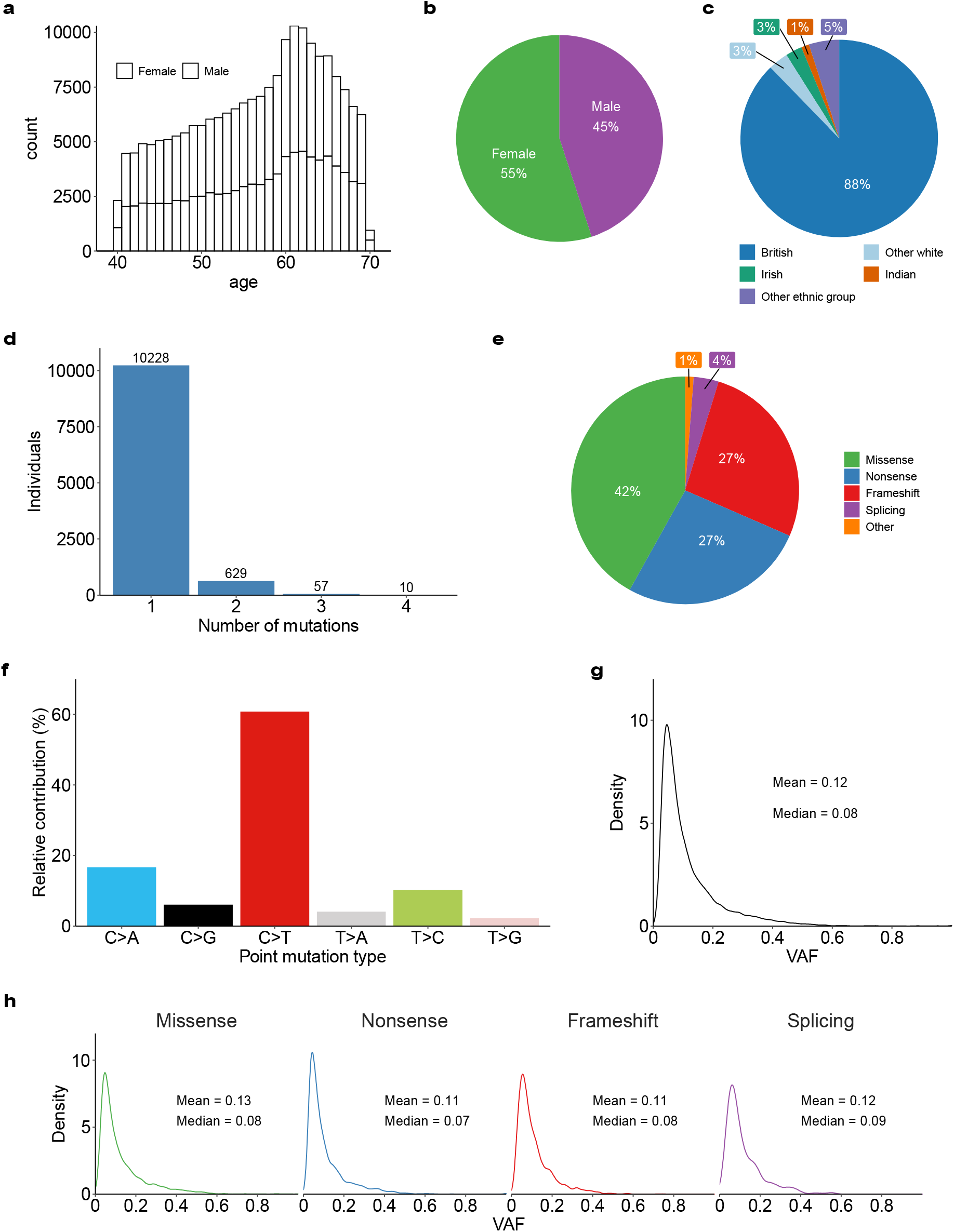
Characterization of CH in the UK Biobank. **a**, Histogram stratified by sex showing the age-distribution of individuals in the UKB cohort (n=200,453). **b**, Overall percentage of females and males in the UKB cohort. **c**, Percentage of the most common self-reported ethnic groups in the UKB cohort. Ethnic groups with a frequency lower than 1% were grouped under the “Other ethnic group” category. **d**, Number of individuals with 1, 2, 3, and 4 somatic mutations. More than 90% of individuals with CH had only one driver mutation identified. **e**, Percentages of different CH mutation types identified. **f**, Relative prevalence of each of the six base substitution types amongst the identified CH mutations. **g**, Density plot showing the variant allele fraction (VAF) distribution of all CH somatic mutations. **h**, Density plot showing similar VAF distribution for different mutation types. Mean and median are indicated for g and h.

**Extended Data Fig. 2:**
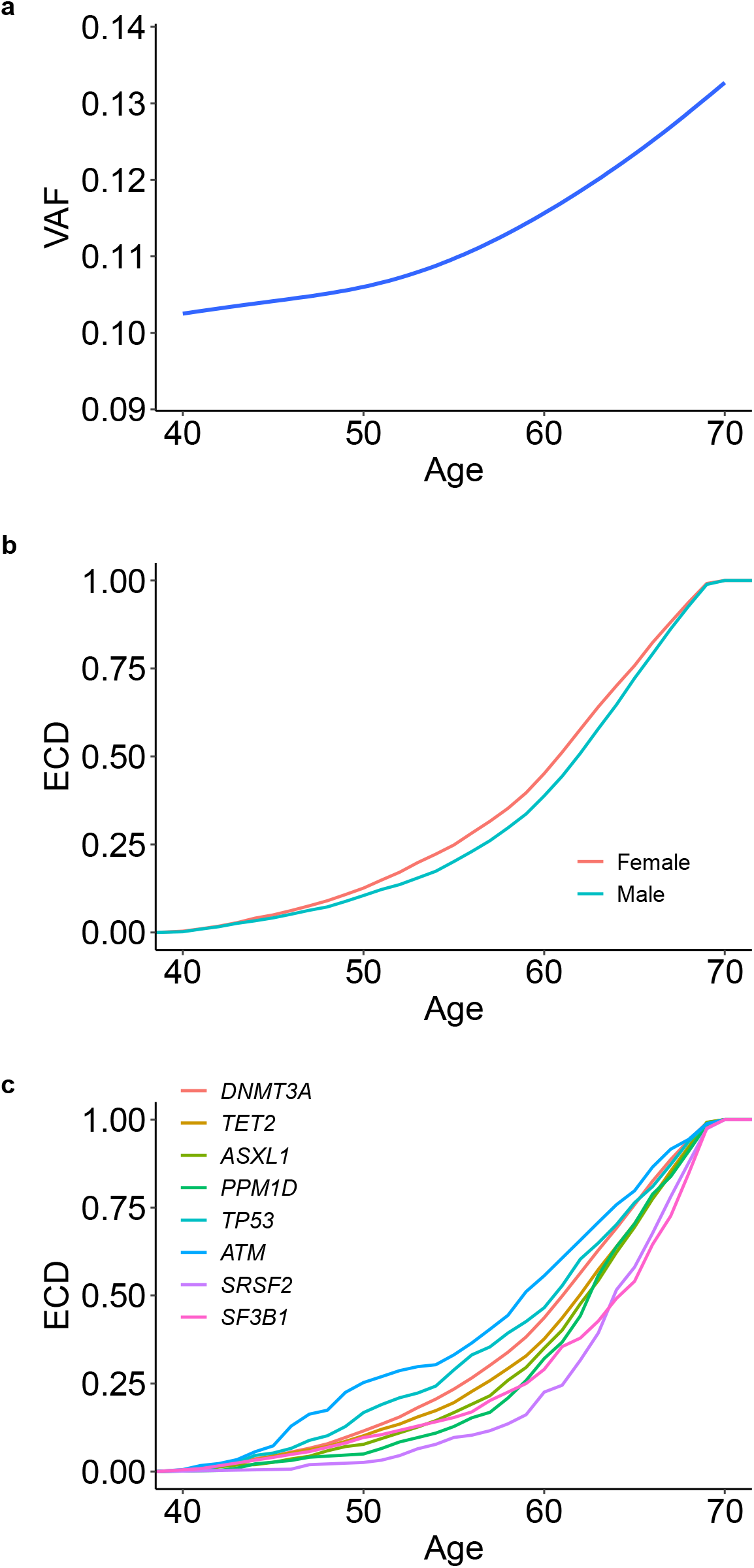
Age-distribution of CH by clone size, sex, and mutant gene. **a**, Clone size, estimated by the variant allele fraction (VAF), increases with age. The blue line represents the smoothed model fitted to a generalized additive model and the shadow represents the 95% confidence interval. **b**, Empirical cumulative distribution (ECD) of the age of individuals with CH stratified by sex. CH was observed one year earlier in females than in males (median 61 versus 62 years; *P*=1.6 × 10^−4^, two-sided pairwise Wilcoxon rank sum test). **c**, ECD of the age of individuals with CH stratified by the eight most common driver genes. Compared to *DNMT3A*, mutations in *ATM* were observed 3 years earlier (*P*=7.2×10^−4^), while mutations in *ASXL1, PPM1D, SRSF2*, and *SF3B1* and were observed 1 (*P*=2.7×10^−8^), 1 (*P*=8.5×10^−6^), 2 (*P*=5.7×10^−10^), and 3 (*P*=6.5×10^−6^) years later, respectively. Differences were calculated using a pairwise Wilcoxon rank sum test.

**Extended Data Fig. 3:**
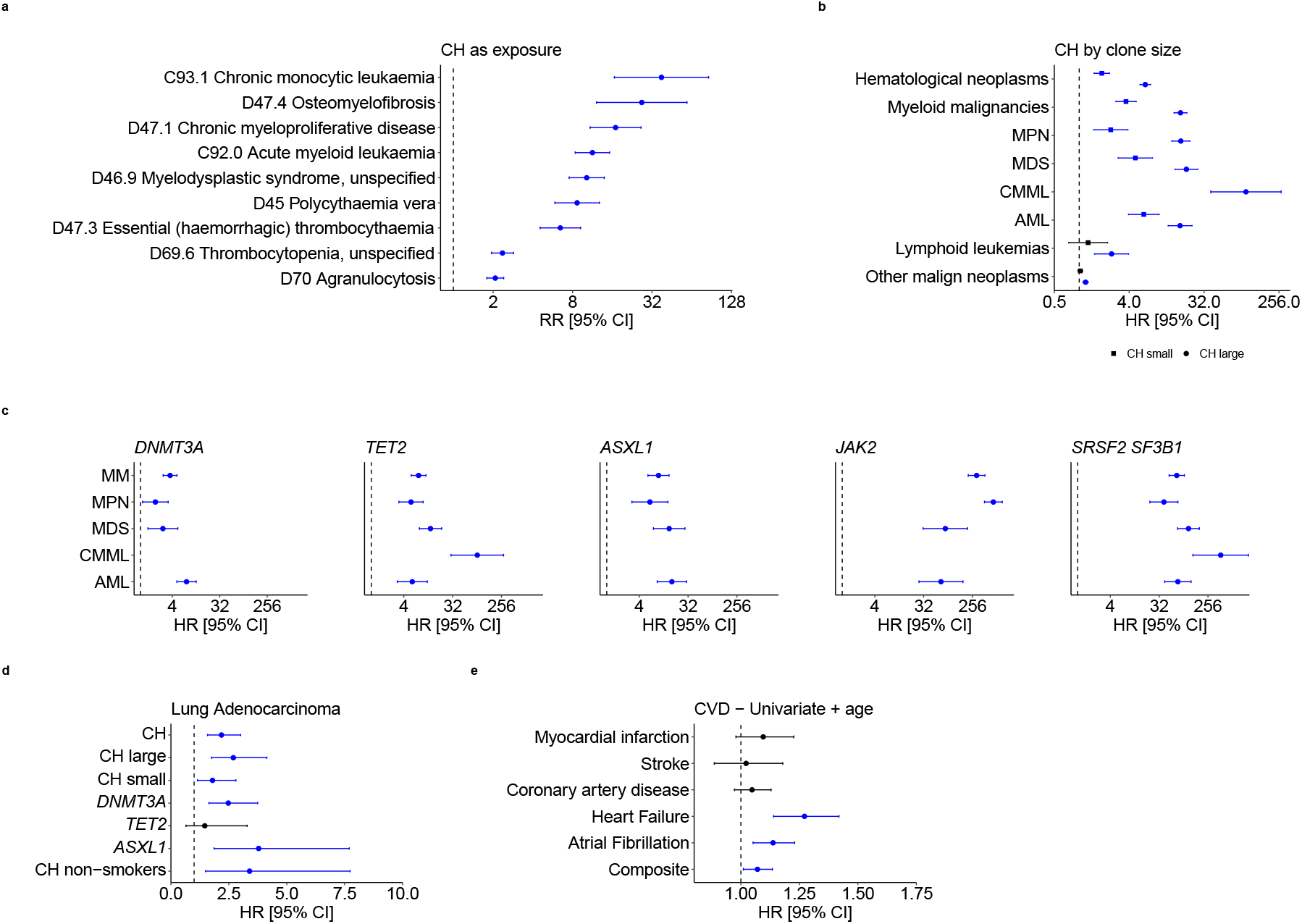
Associations between CH and diseases. **a**, Association analysis of CH with International Classification of Diseases version-10 (ICD-10) disease codes, showing the risk ratios (RRs) for ICD-10 codes with CH as exposure. Only ICD-10 codes with false discovery rate (FDR)<10^−10^ are represented. Error bars represent 95% confidence intervals (CIs). **b-c**, Forest plots showing the hazard ratios (HRs) from Cox proportional-hazards models for association with subsequent hematological and other malignant neoplasms for CH with small and large clones (b) and CH driven by mutations in specific genes (c). **d**, Forest plot showing the HRs for subsequent/incident lung adenocarcinoma for overall CH, CH with large and small clones, and CH with *DNMT3A, TET2*, and *ASXL1* mutations, as well as for overall CH restricting to only self-reported “never-smokers”. **e**, HRs for subsequent/incident cardiovascular disease (CVD) conditions after CH at baseline using a bivariable model containing age as the only covariate. For b-e, HR markers with *P*-value<0.05 are depicted in blue. Error bars represent 95% CIs. Numerical values for RRs/HRs, 95% CIs, and *P*-values are reported in Supplementary Tables 9—12.

**Extended Data Fig. 4:**
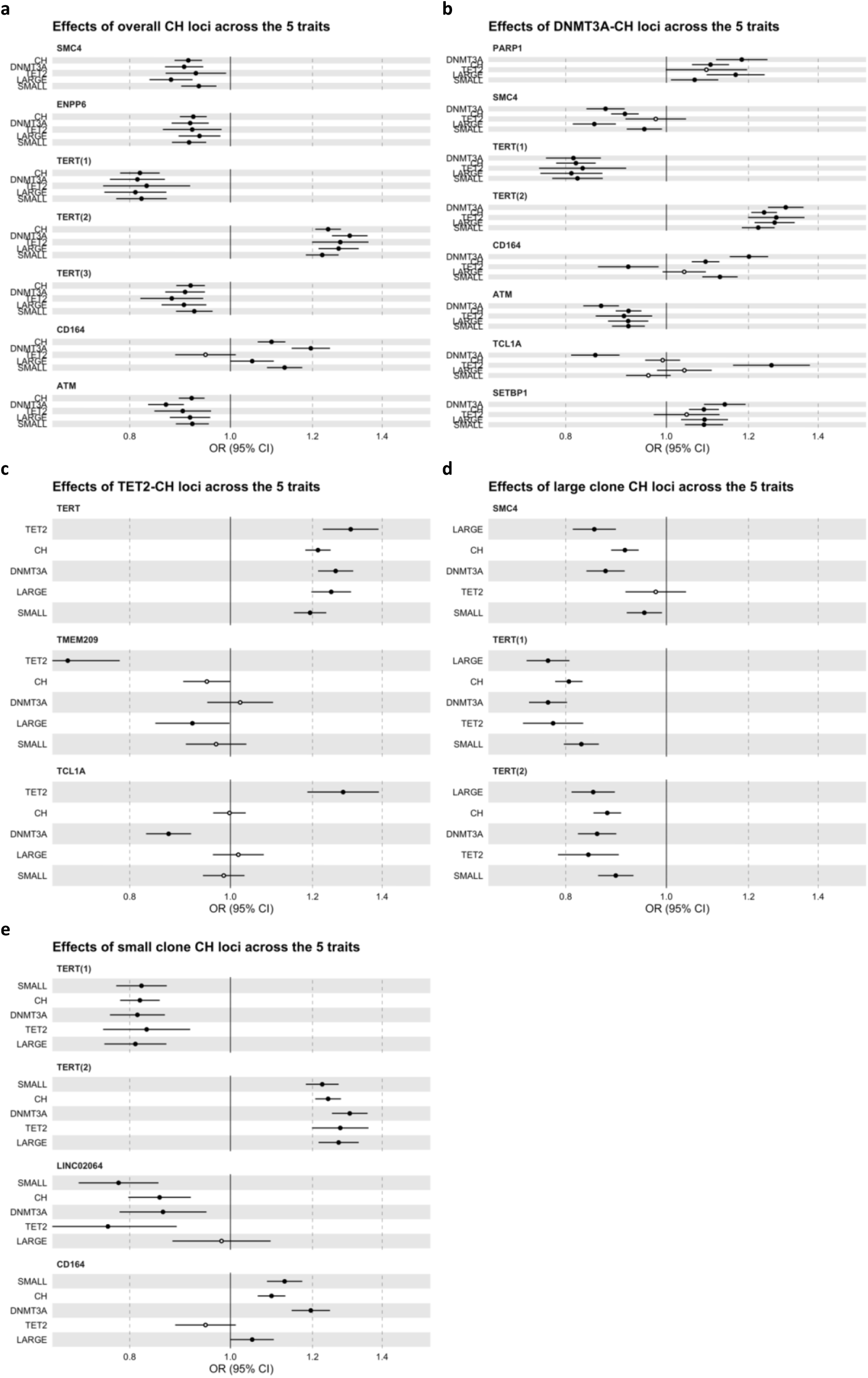
Heterogeneity of lead GWAS variants across five CH traits. Forest plots with odds ratios (ORs) and 95% confidence intervals (CIs) based on data from Supplementary Tables **a**, 16, **b**, 18, **c**, 19, **d**, 20, and **e**, 21. Results for lead variants identified at genome-wide significance (*P*<5×10^−8^) for each CH trait (**a**, overall CH, **b** *DNMT3A*-CH, **c** *TET2*-CH, **d** large clone CH, and **e**, small clone CH) are plotted alongside results for the same lead variants in the four other genome-wide association analyses conducted.

